# Racial and Ethnic Differences in Sodium Sources and Sodium Reduction Behaviors among US Adults: NHANES 2017-2020 pre pandemic

**DOI:** 10.1101/2024.08.27.24312681

**Authors:** Jessica Cheng, Anne N Thorndike, Stella Yi

## Abstract

**Background:** Nearly all US adults exceed sodium recommendations, which increases cardiovascular risk. Understanding racial and ethnic differences in sodium sources and behaviors could lead to nuanced public health messaging, dietary interventions, and clinical guidance to more equitably achieve population-level sodium reduction.

**Methods:** Using National Health and Nutrition Examination Survey 2017-2020 pre-pandemic data, racial and ethnic differences in sodium sources and sodium-related behaviors (e.g., salt use at the table and in food preparation, doctor advice to reduce sodium, attempts to reduce sodium, and label reading) were assessed using weighted chi-square. Given the nutrient database’s assumption that rice is salted may be inappropriate for some ethnic groups, we conducted a secondary analysis altering this assumption.

**Results:** Pizza, soup, and chicken were top sources of sodium across racial and ethnic groups. For Asian Americans, 4 top sources were unique (e.g., soy-based condiments). Black adults reported the highest rates of reducing sodium (67% vs. 44% among White adults) and receiving physician sodium reduction advice (35% vs.18% among Asian Americans). Asian Americans were the most likely to frequently use salt during food preparation (66% vs. Other Race adults 32%) but reported not using salt at the table (43% vs. 23% among Other Race adults). Assuming rice is unsalted reduces Asian American sodium intake estimates by ∼325 mg/day.

**Conclusions:** While product reformulation targets and front-of-pack nutrition labeling may help reduce sodium intake across groups, to equitably address sodium intake, culturally appropriate advice on sources of sodium and salt usage may be needed, particularly for Asian Americans.

**Clinical Perspective:** *What Is New?:*

- Among the top 10 sources of Asian American adult sodium intake, 4 sources accounted for over 14% of sodium intake and were not shared with other racial and ethnic groups (e.g., soy-based condiments, fish, fried rice/lo mein, and stir-fry and soy-based sauce mixtures) while few unique sources were identified for other racial and ethnic groups.
- Black adults reported the highest rates of reducing sodium and receiving physician sodium reduction advice, and Asian Americans were the most likely to frequently use salt during food prep but least likely to use salt at the table.
- Compared to estimates derived under the Food and Nutrient Database for Dietary Studies (FNDDS) assumption that rice is salted, applying the assumption that rice is unsalted lowers Asian American sodium intake estimates by about 325 mg/day.

*What Are the Clinical Implications?:*

- Because of different top sources of sodium, public health messaging, clinical guidance, and product reformulation efforts will need to focus on multiple food products used across racial and ethnic groups.
- Tailored advice on salt usage and culturally and linguistically appropriate patient education may support physician advice to reduce sodium while front of pack nutrition labelling may improve label reading across racial and ethnic groups.
- To improve sodium surveillance in the US, both salted and unsalted rice should be included in dietary databases and assessment of sodium intake using a less biased measure (e.g., 24-hour urinary sodium excretion) should be considered.

## Introduction

The average sodium intake of US adults (3,346 mg/day)^1^ is higher than the 2,300 mg/day limit recommended by the Dietary Guidelines for Americans.^2^ Sodium intake has remained elevated^3, 4^ despite Healthy People 2020 and 2030 goals^5, 6^ and voluntary product reformulation targets.^3, 4, 7^ As cardiovascular disease is the top cause of death,^8^ and decreasing sodium intake has been shown to reduce cardiovascular morbidity and mortality,^9–11^ both through hypertension and independent of it,^12, 13^ sodium reduction is an important national and international health priority.^6, 14^ As there are racial and ethnic differences in hypertension^15^ and CVD,^16, 17^ it is valuable to understand differences in the top sources of sodium and sodium reduction behaviors by race and ethnicity.

In fact, an earlier analysis of NHANES 2011-2012 showed 6 of the top 10 sodium sources specific to Asian Americans (e.g., stir-fry and soy-based sauces) were not among the top 10 sources for any other racial and ethnic group.^19^ The analysis also revealed differences by race and ethnicity in salt usage with Asian Americans adding salt more frequently during food preparation than other groups but less frequently adding salt at the table.^19^ Updated estimates of racial and ethnic specific sodium sources and sodium behaviors are needed given demographic shifts, for example, increases in the Asian American population^20^ and count of those born outside the US.^21^

If population-level sodium reduction is to be achieved, researchers, healthcare practitioners, and policymakers must focus on diverse groups and their sodium-related behaviors and consumption. ^22–24^ The National Institute of Minority Health and Health Disparities (NIMHD) Research Framework^25^ can help guide interpretation of differences in sodium sources and behaviors and the implications for health equity work. For example, behavioral influences on health outcomes at the individual level (e.g., differences in sodium sources) can have implications for societal level behavioral strategies (e.g., culturally tailoring public health campaigns, community-based health interventions, and clinical practice recommendations). Such tailoring efforts are worthwhile as evidence shows greater effectiveness of culturally tailored interventions compared to generic interventions.^22–24^

The purpose of this analysis was to describe the top 10 sources of sodium for each racial and ethnic group separately using the most up-to-date data (i.e., NHANES 2017-2020 pre-pandemic). To further inform culturally appropriate targets for sodium reduction, we assessed racial and ethnic differences in individual and interpersonal behavioral factors that could be modified to decrease sodium intake, including nutrition label reading for sodium content, physician advice to reduce sodium, salt usage at the table and in food preparation, personal efforts to reduce sodium, and type of salt used.

## Methods

The analytical code used to generate results is publicly available and can be accessed at Open Science Framework (DOI 10.17605/OSF.IO/ER4V9). JC had full access to all the data and takes responsibility for its integrity and the data analysis. The National Health and Nutrition Examination Survey (NHANES) 2017-2020 pre-pandemic public use dataset was used for this analysis and is representative of the non-institutionalized US population. Data were collected from 2017 to March 2020. Briefly, NHANES utilizes a complex, stratified, multistage probability cluster sampling method. Detailed information on sampling, data collection procedures, and response rates is described elsewhere.^26, 27^

All adults (≥18 years) were included in this analysis, including pregnant and lactating women, as the sodium recommendations are the same as in the general population unless there is a specific medical indication.^28^ Race and ethnicity were self-reported. Because of heterogeneity within the Hispanic group, we chose to disaggregate the data. Participants were grouped into “Mexican American”, “Other Hispanic”, “non-Hispanic Asian”, “non-Hispanic Black”, “Other Race including multi-racial”, and “non-Hispanic White” categories hereafter referred to as Mexican American, Other Hispanic, Asian American, Black, Other Race, and White.

### Sodium Sources

Dietary information was collected using the National Cancer Institute’s (NCI) Automated Self-Administered (ASA-24) dietary recall system.^29^ The first recall, collected in person with a trained interviewer, was used for this analysis as has been done in prior analyses.^18, 19, 30–32^ The United States Department of Agriculture Food and Nutrient Database for Dietary Studies (FNDDS) is used by ASA-24 to determine the sodium intake of reported foods and beverages. Foods and beverages were grouped into 169 categories according to the What We Eat in America (WWEIA) classification.^33^ This classification system has been used in other studies reporting sources of sodium.^18, 19^ Sodium intake from dietary supplements was not included as very little sodium comes from these sources.^34^

### Sodium Reduction Behaviors

Participants self-reported the frequency of using food labels to check sodium content (i.e., Always, most of the time, sometimes, rarely, never, never seen). Because of small numbers, ‘never’ and ‘never seen’ were collapsed into a single category. Participants responded yes or no to whether their doctor has told them to reduce salt in their diet in the past 12 months and whether they are now reducing salt.

Participants self-reported the type of salt used at the table (i.e., ordinary salt [includes regular iodized salt, sea salt and seasoning salts made with regular salt], lite salt, salt substitute, doesn’t use or add salt at the table). Because of small numbers, ‘lite salt’ and ‘salt substitute’ were collapsed into a single category. If using salt at the table, participants self-reported the frequency of salt use (i.e., rarely, occasionally, very often). Participants also self-reported salt use in food preparation (i.e., never, rarely, occasionally, very often).

### Analysis Plan

We descriptively present the top 10 sources of sodium by racial and ethnic group using the National Cancer Institute’s population proportion method.^35^ Briefly, sodium intake for each food category was summed across individuals and divided by the sum of total daily sodium intake for all individuals. Differences in sodium behaviors by race and ethnicity were examined using the adjusted Wald F chi-square statistic.

Data analysis procedures were adapted from NHANES example programs.^36^ All procedures (i.e., *proc ratio, proc crosstabs*) were survey-weighted (e.g., WTDRD1PP, WTINTPRP, WTMECPRP) and accounted for the complex sampling design (e.g., SDMVSTRA SDMVPSU) using Taylor Series Linearization with appropriate *subpopx* statements. *P<0.05* was considered statistically significant. As recommended, Korn-Graubard corrected confidence intervals are presented and National Center for Health Statistics (NCHS) Data Presentation Standards for Proportions applied using the *KG_macro*.^37, 38^ Unstable estimates are presented in tables with a note.

We ran a sensitivity analysis. NHANES pulls nutrient information from FNDDS. FNDDS assumes foods in the WWEIA category ‘rice’ include iodized table salt as an ingredient. However, there is limited ethnographic evidence to demonstrate all Asian ethnic groups (e.g., Chinese, Filipino, Vietnamese, Asian Indian) salt water when cooking rice. Therefore, to see the effect of this assumption on top sodium sources, we re-ran the analysis assuming no salt added to food codes in the rice category except for yellow rice. Sodium estimates for WWEIA category ‘fried rice and lo/chow mein’ (e.g., ‘rice, fried, with pork’) and ‘rice mixed dishes’ (e.g., ‘congee, with meat, poultry, and/or seafood’) were not altered.

For example, 100 grams of ‘Rice, white, cooked, no added fat’ has 246 mg of sodium. As there is 38800 mg of sodium for every 100 grams of ‘Salt, table, iodized,’ and cooked rice has little sodium inherently (i.e., 1 mg per 100 grams),^39^ over 99% of the sodium in ‘Rice, white, cooked, no added fat’ was removed. Nutrient information for each food code in the rice category is available at FoodData Central^40^ and provided in Supplemental Table 1.

**Table 1:** Weighted Percent Contribution and 95% Confidence Intervals* of the Top 10 Sources of Sodium Intake among US Adults by Race thnicity, NHANES 2017-2020 pre-pandemic.

| Rank | Mexican American (N=952) |  | Other Hispanic (N=819) |  | Other Hispanic Sensitivity Analysis† (N=819) |  | Asian American (N=877) |  | Asian American Sensitivity Analysis† (N=877) |  | Black (N=2153) |  | Other Race (N=397) |  | White (N=2893) |  |
| --- | --- | --- | --- | --- | --- | --- | --- | --- | --- | --- | --- | --- | --- | --- | --- | --- |
|  | Source | % (95% CI) | Source | % (95% CI) | Source | % (95% CI) | Source | % (95% CI) | Source | % (95% CI) | Source | % (95% CI) | Source | % (95% CI) | Source | % (95% CI) |
| 1 | Burritos and tacos | 12.3 (9.7, 15.2) | Soups | 6.6 (4.6, 9.1) | Soups | 6.8 (4.7, 9.4) | Soups | 11.9 (9.7, 14.1) | Soups | 13.0 (10.7, 15.6) | Chicken, whole pieces | 7.7 (6.6, 8.9) | Chicken, whole pieces | 5.3 (3.4, 7.9) | Pizza | 5.30 (4.5, 6.1) |
| 2 | Soups | 5.7 (4.4, 7.3) | Chicken, whole pieces | 5.4 (3.2, 8.3) | Chicken, whole pieces | 5.5 (3.3, 8.6) | Rice | 8.6 (6.6, 10.5) | Soy-based condiment s<sup>§</sup> | 5.6 (2.8, 10.0) | Pizza | 5.2 (4.0, 6.7) | Pasta mixed dishes, excludes macaroni and cheese | 5.3 (2.8, 9.0) | Cold cuts and cured meats | 42 (3.3, 5.2) |
| 3 | Pizza | 5.1 (3.4, 7.4) | Pizza | 5.1 (3.4, 7.2) | Pizza | 5.2 (3.5, 7.4) | Soy-based condiment s<sup>§</sup> | 5.2 (2.1, 8.2) | Yeast breads | 4.7 (3.8, 5.8) | Cold cuts and cured meats | 3.2 (2.2, 4.6) | Pizza | 3.5 (2.2, 5.3) | Soups | 4.2 (3.4, 5.0) |
| 4 | Other Mexican mixed dishes | 4.2 (3.1, 5.6) | Burritos and tacos | 4.2 (2.7, 6.2) | Burritos and tacos | 4.4 (2.8, 6.4) | Yeast breads | 4.3 (3.4, 5.2) | Chicken, whole pieces | 4.4 (3.3, 5.8) | Burgers | 3.0 (2.2, 4.1) | Meat mixed dishes<sup>†</sup> | 3.4 (0.8, 9.2) | Chicken, whole pieces | 3.2 (2.6, 3.8) |
| 5 | Chicken, whole pieces |  | Rice | 3.3 (2.3, 4.6) | Pasta mixed dishes, excludes macaroni and | 2.9 (2.0, 4.2) | Chicken, whole pieces | 4.0 (2.9, 5.2) | Fish | 3.8 (1.7, 7.2) | Soups | 3.0 (2.2, 3.9) | Cold cuts and cured meats | 3.3 (1.7, 5.6) | Meat mixed dishes | 3.1 (2.4, 3.9) |
| 6 | Tomato-based condiments | 2.9 (2.3, 3.7) | Pasta mixed dishes, excludes macaroni and cheese | 2.8 (1.9, 4.0) | Eggs and omelets | 2.7 (2.1, 3.4) | Fish | 3.5 (1.2, 5.7) | Pizza | 3.6 (2.5, 5.0) | Pasta mixed dishes, exclude s macaroni and cheese | 2.7 (2.2, 3.3) | Cheese | 3.2 (1.9, 4.9) | Cheese | 3.0 (2.6, 3.6) |
| 7 | Eggs and omelets | 2.6 (2.3, 3.0) | Eggs and omelets | 2.6 (2.0, 3.3) | Yeast breads | 2.6 (1.9, 3.5) | Pizza | 3.3 (2.2, 4.5) | Fried rice and lo/chow mein | 2.5 (1.5, 3.9) | Burritos and tacos | 2.6 (1.8, 3.8) | Soups | 3.1 (1.8, 5.1) | Yeast breads | 3.0 (2.4, 3.6) |
| 8 | Cold cuts and cured meats | 2.6<br>(1.7, 3.8) | Yeast breads | 2.6<br>(1.9, 3.4) | Deli and cured meat sandwiches | 2.6<br>(1.4, 4.3) | Fried rice and lo/chow mein | 2.3<br>(1.2, 3.3) | Stir-fry and soy-based sauce mixtures | 2.4<br>(1.4, 3.8) | Cheese | 2.3<br>(2.0, 2.7) | Egg/breakfast sandwiches<sup>‡</sup> | 2.9<br>(0.9, 6.6) | Pasta mixed dishes, excludes macaroni and cheese | 2.9<br>(2.4, 3.5) |
| 9 | Beans, peas, legumes | 2.5<br>(1.7, 3.5) | Deli and cured meat sandwiches | 2.5<br>(1.4, 4.2) | Beans, peas, legumes | 2.5<br>(1.8, 3.3) | Stir-fry and soy-based sauce mixtures | 2.2<br>(1.2, 3.2) | Meat mixed dishes | 2.2<br>(1.4, 3.2) | Yeast breads | 2.3<br>(1.9, 2.7) | Tomato-based condiments | 2.6<br>(1.3, 4.8) | Deli and cured meat sandwiches | 2.6<br>(2.0, 3.3) |
| 10 | Egg/breakfast sandwiches | 2.3<br>(1.7, 3.0) | Beans, peas, legumes | 2.4<br>(1.8, 3.2) | Burgers | 2.4<br>(1.8, 3.1) | Meat mixed dishes | 2.0<br>(1.2, 2.8) | Burritos and tacos | 2.1<br>(1.3, 3.4) | Chicken patties, nuggets and tenders | 2.0<br>(1.5, 2.7) | Burgers | 2.6<br>(1.3, 4.6) | Burgers | 2.5<br>(2.0, 3.2) |
| **Cumulative %** |  | 43.9 |  | 37.4 |  | 37.6 |  | 47.2 |  | 44.3 |  | 34.1 |  | 35.1 |  | 33.8 |

\*Confidence intervals were calculated using the Korn-Graubard method.
<sup>†</sup> A sensitivity analysis was run for Asian Americans whereby food codes under WWEIA Category Number: 4002 corresponding to WWEIA Category Description: Rice (food codes: 56205000, 56205001, 56205002, 56205004, 56205006, 56205007, 56205008, 56205011, 56205012, 56205014, 56205016, 56205017, 56205018, 56205060, 56205070, 56205101, 56205190, 56205205, 56205210, 56205215, 56205300, 56205310, 56205320, 56205330, 56205340, 56205350, and 56205410) were assumed to be unsalted except yellow rice (food codes: 56205130, 56205150, and 56205170).
<sup>‡</sup>Estimates may be unstable because confidence interval width CIW>5 and relative confidence interval width RCIW>130.
<sup>§</sup>WWEIA category 8404 "Soy-based condiments" includes fish and oyster sauce. Although these products are not generally soy-based, they were not recategorized for the analysis. However, food code 89901040 "Soy-based sauce, for use with vegetables" was recategorized from WWEIA category 9999 "Uncategorized" to WWEIA category 8404 "Soy-based condiments".

Additionally, we used day 1 and day 2 dietary recalls and the Simulating Intake of Micronutrients for Policy Learning and Engagement (SIMPLE) macro,^41^ after generating balanced repeated replicates,^42^ to describe the mean and distribution of sodium intake by race and ethnicity when varying the assumption that rice is salted. Estimates were adjusted for a weekend/weekday indicator. Technical details of the SIMPLE macro, which implements the NCI method for estimating usual dietary intake, can be found elsewhere.^41, 43^ All analyses were run in SAS 9.4 (Cary, NC) and SAS-callable SUDAAN 11.0.4.

## Results

### Sodium Sources

The racial and ethnic-specific top 10 sources of sodium intake account for about one-third (for White adults) to half (for Asian Americans) of sodium intake (Table 1). Soups, pizza, and chicken, whole pieces were among the top 10 sources for every racial and ethnic group. However, the ranking of these items and percentage of sodium intake they represented differed. For example, soups made up 12% of Asian American sodium intake compared to 3% of intake among Other Race adults. Other food categories (e.g., yeast breads, cold cuts, and burritos and tacos) were common across most groups.

For Asian Americans, 4 of the top 10 sources were unique to their group only (i.e., soy-based condiments, fish, fried rice and lo/chow mein, and stir-fry and soy-based sauce mixtures). For Mexican Americans, other Mexican mixed dishes was a unique top source of sodium, and for Black adults, chicken patties, nuggets, and tenders was a unique top source. Mexican Americans and Other Hispanic adults had 6 out of 10 top sources in common.

### Sodium Reduction Behaviors

About one-third of participants across racial and ethnic groups indicated rarely or never using nutrition labels or never having seen the labels (Table 2) (p=0.05). Across racial and ethnic groups, about half of all US adults indicated they were now reducing sodium in their diet with the lowest rate among White adults (44.0%) and the highest rate among Black adults (66.5%) (p<0.001). There were racial and ethnic differences in self-reported doctor advice to reduce sodium (p<0.01). For example, 18% Asian Americans reported their doctor talked to them about reducing sodium compared to 35% of Black adults.

**Table 2:**
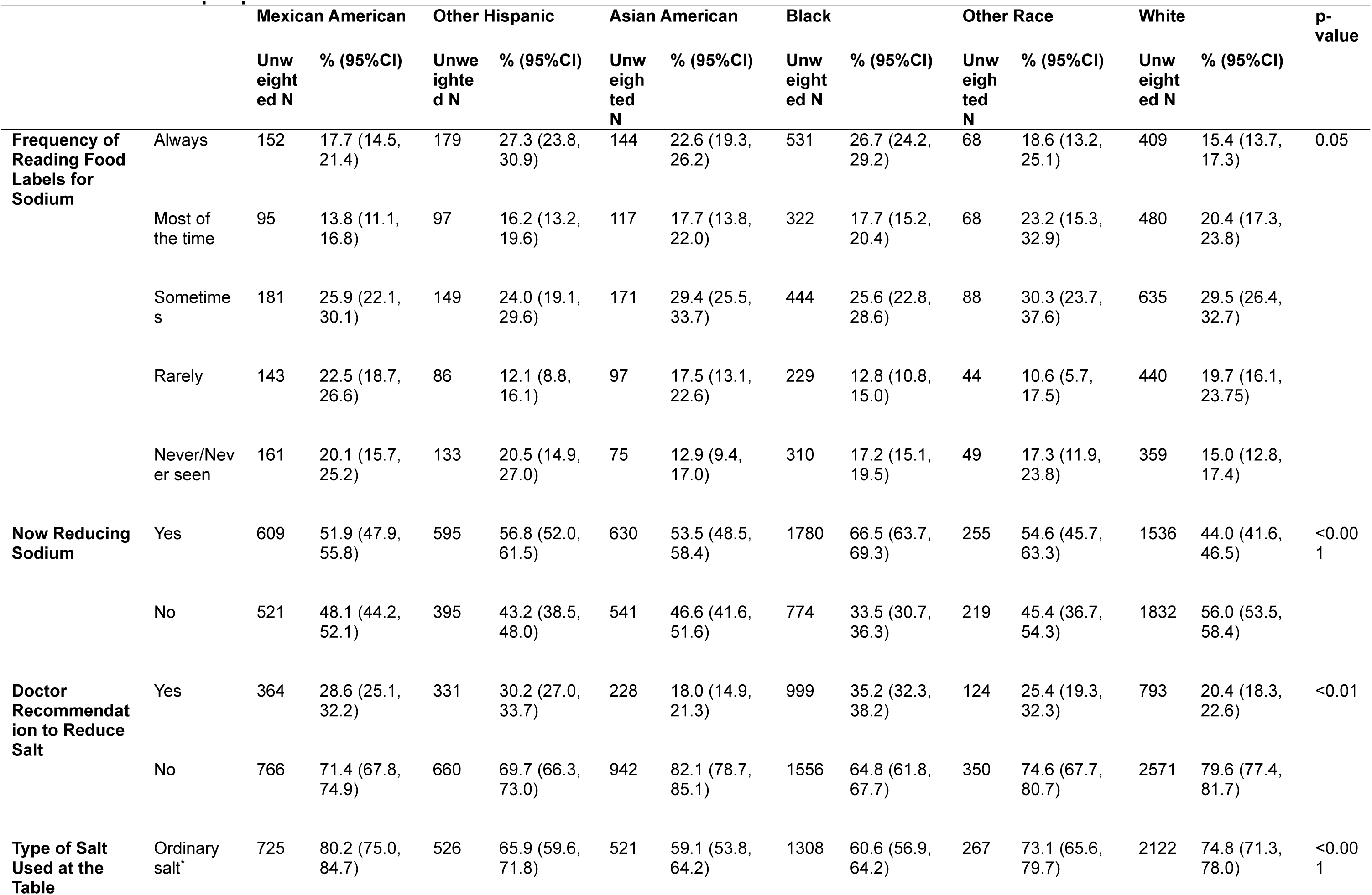
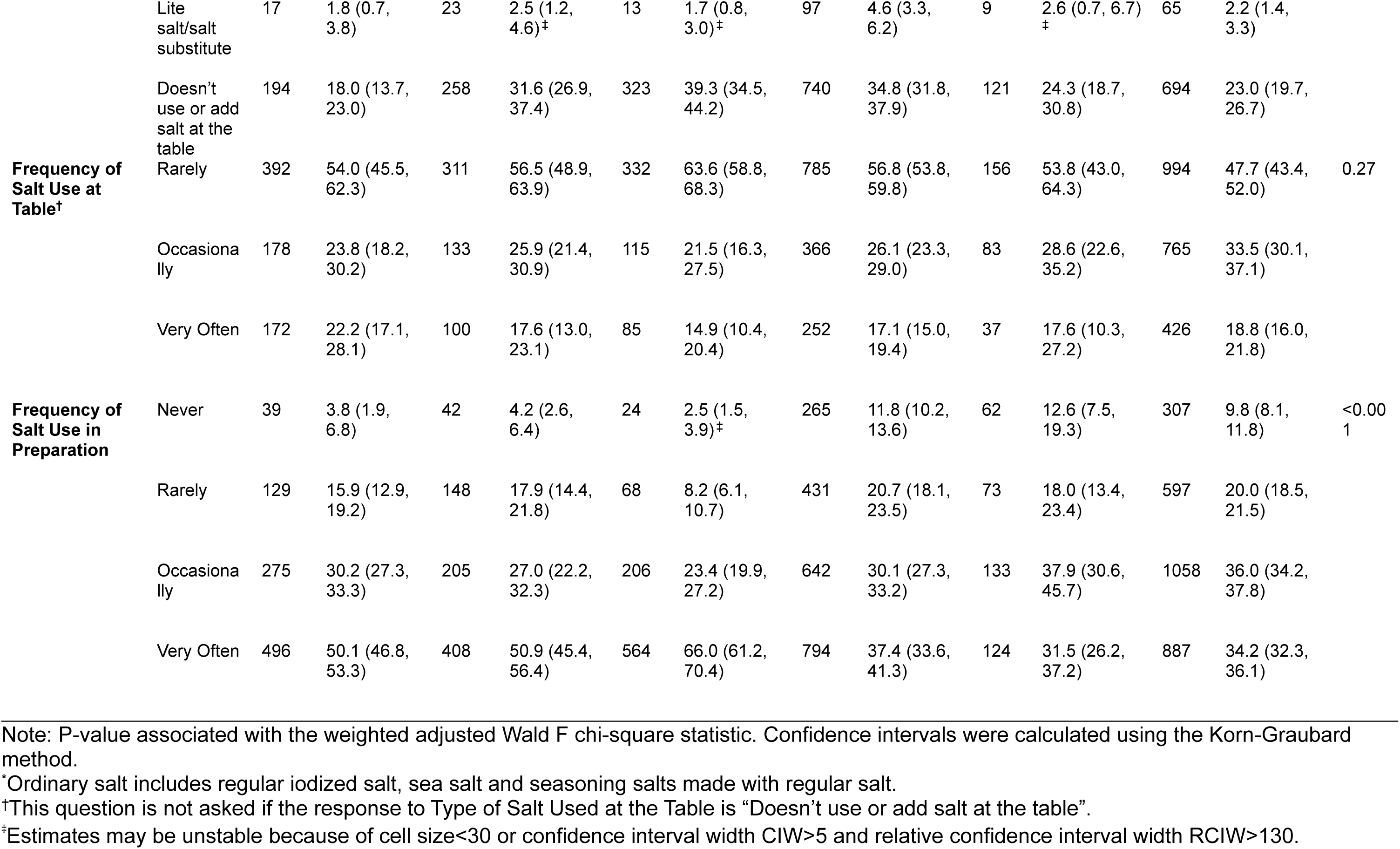
Weighted Frequency and 95% Confidence Intervals of Sodium Reduction Behaviors among US Adults by Race and Ethnicity, NES 2017-2020 pre-pandemic.

There were also differences in salt use at the table (p<0.001). While most adults use salt at the table, 23% of Other Race adults indicated not using salt at the table compared to 43% of Asian Americans. Among those who use salt at the table, almost all adults used regular salt as opposed to lite salt or a salt substitute with the percentage of adults reporting salt usage at the table “very often” ranging from 14.9% among Asian Americans to 22.2% among Mexican Americans (p=0.27). However, there were differences in adding salt during cooking (p<0.001), with the percentage reporting salt usage in food preparation “very often” ranging from 31.5% of Other Race adults to 66.0% among Asian Americans.

### Sensitivity Analysis

Rice was the second most important source of sodium for Asian Americans, accounting for nearly 9% of intake. For Other Hispanic adults, rice was the 5^th^ top source of sodium accounting for 3.3% of intake. In a sensitivity analysis assuming no salt in rice, rice was no longer a top source of sodium, accounting for <0.5% of sodium for both Asian Americans and Other Hispanic adults (Table 1).

Varying the assumption of salt added to rice appeared to have little effect on sodium estimates for most groups (Figure 1 and Supplemental Table 2); however, for Other Hispanic adults, average sodium intake decreased by 118 mg/day (mean [SD]: 3232 [92.5] to 3114 [97.7] mg/day) and for Asian Americans by 323 mg/day (3500 [60.7] vs. 3177 [64.6] mg/day). Although most adults had excessive sodium intake (>2300 mg/day) (Supplemental Table 2 and Supplemental Figure 1), varying the assumption of salt in rice decreased the percentage with excessive intake from 83.1% to 79.5% for Other Hispanic adults and from 89.0% to 81.4% for Asian Americans.

**Figure 1:**
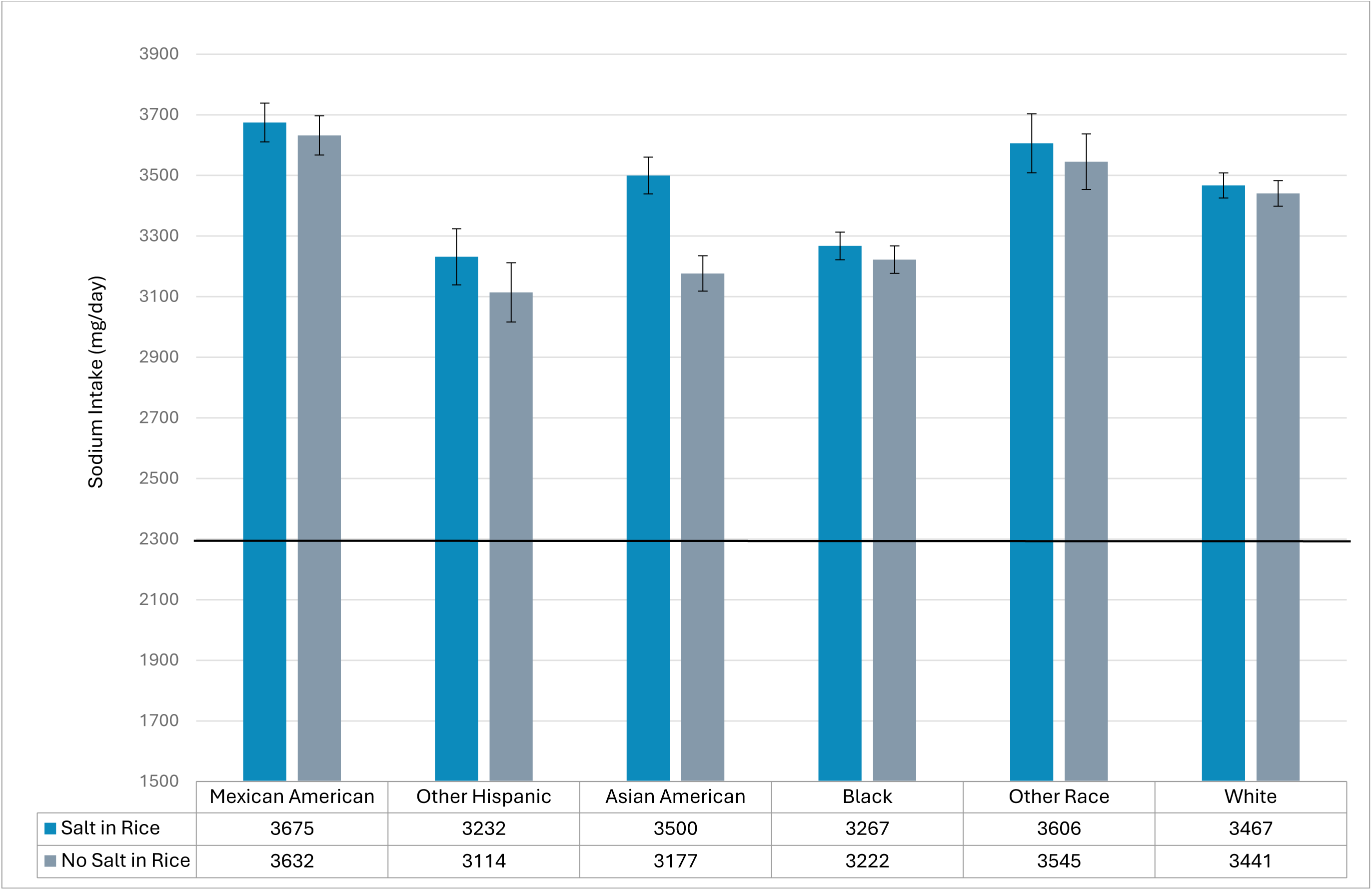
Mean and Standard Error of US Adult Sodium Intake by Race & Ethnicity under Varying Assumptions about Salt in Rice, NES 2017-2020 pre-pandemic. Note: The 2020-2025 Dietary Guidelines for Americans recommend sodium intake below 2300 mg/day which is indicated by the black horizontal line on the figure. “Rice” includes food codes: 56205000, 56205001, 56205002, 56205004, 56205006, 56205007, 56205008, 56205011, 56205012, 56205014, 56205016, 56205017, 56205018, 56205060, 56205070, 56205101, 56205130, 56205150, 56205170, 56205190, 56205205, 56205210, 56205215, 56205300, 56205310, 56205320, 56205330, 56205340, 56205350, and 56205410. Salt in Rice: The sodium content was not altered for any of the food codes in the “rice” category. No salt in Rice: Sodium due to salt was removed from all foods codes in the “rice” category except for yellow rice (foods codes: 56205130, 56205150, and 56205170).

Additionally, the estimates suggested heterogeneity of sodium intake among Hispanic adults. Mean sodium consumption was highest among Mexican Americans while Other Hispanic adults had the lowest sodium intake. Of note, the Other Race group, whose intake is not always enumerated, had the second highest sodium intake.

## Discussion

Results showed both similarities and differences in sources of sodium and sodium reduction behaviors between racial and ethnic groups. These findings have implications for health equity as sodium reduction strategies may benefit groups differently. Although about half of all US adults indicated they were reducing sodium in their diet, average sodium intake exceeded recommended levels for all racial and ethnic groups. Thus, in alignment with the NIMHD Research Framework,^25^ continued efforts to reduce sodium at the societal, community, interpersonal, and individual levels are needed in order to aid adults in their efforts to reduce sodium equitably. These efforts will need to draw on the unique strengths and address the unique barriers to sodium reduction in different subgroups. Below we discuss how similarities and differences in individual and interpersonal sodium sources and sodium-related behaviors relate to US sodium reduction strategies at multiple levels.

Our findings on the top sources of sodium in the US diet were similar to earlier reports. For example, all reports identified pizza and soups as top sources across racial and ethnic groups^18, 19, 30^ with other sources shared across multiple reports and groups (e.g., yeast breads). Therefore, one strategy to reduce population level sodium intake would be to reformulate common foods. Simulation work in the US suggests changes by industry to the food supply would make a meaningful impact on sodium intake.^44–48^ Although the impact would vary across racial and ethnic groups and socioeconomic levels, decreases in sodium intake would be expected for all with the largest impacts among some of the least advantaged groups.^44–48^

To address sodium levels in common foods such as pizza and bread products, the National Salt and Sugar Reduction Initiative,^49^ supported by the Food and Drug Administration (FDA),^7, 50^ has set voluntary standards for industry to meet. While reductions in sodium have been observed in the food supply since the introduction of these targets,^51^ progress on voluntary sodium reduction has slowed.^52^ Besides finding additional ways to incentivize industry to meet these targets, FDA can take other steps to support reformulation efforts. Product reformulation has been successfully implemented in several countries in part by utilizing lite salt and salt substitutes.^14, 53^ Lite salt and salt substitutes replace some or all of the sodium chloride in salt with minerals such as potassium, which has beneficial effects on blood pressure.^54^ Thus, the FDA has proposed a rule to amend the ‘standards of identity’ such that salt substitutes may be used in products.^55^ This could help manufacturers meet sodium reduction targets. Continued monitoring of these policies will be needed to ensure uptake among a broaden set of manufacturers and equitable population level effects.

In addition to similarities, we also observed racial and ethnic differences in top sodium sources. Resources that may be provided in primary care, community interventions, or as part of public health campaigns like the American Heart Association’s “Salty Six” infographic focus on breads, pizza, sandwiches, cold cuts, soup, and burritos and tacos as major sources of sodium.^32, 56^ However, as certain items are unique to only a single group (e.g., soy-based condiments for Asian Americans), creating and disseminating versions of these materials^57^ that highlight unique sources may help educators culturally tailor dietary advice.

In addition to targeting foods, another option for reducing sodium is to alter discretionary salt usage by replacing regular salt with lite salt or a salt substitute. Our analysis indicates that currently less than 5% of American adults are utilizing lite salt or salt substitutes despite its availability in US supermarkets.^58^ Recent large randomized controlled trials in Asia and South America provide evidence that discretionary use of sodium potassium salt substitutes may prevent hypertension, reduce blood pressure, and lead to lower rates of stroke, major cardiovascular events, and all-cause mortality.^59–62^ Sodium potassium salt substitutes have an acceptable taste profile,^63–65^ nearly indistinguishable from regular salt.^66^ Indeed, participants in these randomized trials reported near daily use of the substitutes.^61, 67^

However, it has been argued a discretionary salt substitution strategy could not work in the US because of high processed food consumption.^44^ As this strategy has been shown to be effective in Asia and South America, such discretionary salt substitution would likely be culturally acceptable for Asian and Hispanic subgroups in the US. Given over half of Mexican Americans, Other Hispanic adults, and Asian American adults indicate frequent use of salt during cooking and that ethnic minority and immigrant populations in the US report more home cooking than other groups,^68, 69^ discretionary salt substitution may be a viable strategy for lowering sodium consumption for these groups.

While NHANES does not quantify percent contribution to sodium intake of salt added at the table or in food preparation, it is estimated that these sources each account for 5% of total sodium intake with variation by race and ethnicity (e.g., 6.6% and 8.6% of sodium may come from salt added in food preparation for Asian Americans and Hispanic adults, respectively).^34^ This aligns with our finding of racial and ethnic differences in salt use practices. As many of the top 10 sources in this analysis contribute <5% to sodium intake, targeting discretionary salt usage in combination with other sodium reduction strategies may have a sizeable impact on sodium intake.

We found few adults self-reported their doctor advised them to reduce sodium with Black adults the most likely to report having been given advice and Asian Americans the least likely. This is mirrored by provider reports that they advise Asian American patients to reduce sodium intake less frequently than they advise Black patients.^70^ While low rates of physician advice among Asian Americans have been previously reported,^71^ differences between Asian Americans and other groups may underestimate disparities in physician advice as fewer Asian Americans sampled in NHANES completed data collection in a non-English/non-Spanish language than would be expected given that about 1 in 3 have limited English language proficiency^72, 73^ and use of interpreter services in outpatient settings is low.^74^

While increasing physician advice on sodium reduction is needed for all racial and ethnic groups, it will be important to pay attention to obstacles to patient care including cultural discordance between patients and the physicians, nurses, and dietitians who treat them (e.g., over 80% of registered dietitians and registered dietitian nutritionists are White).^75^ However, language discordance between patient and provider may be even more important than cultural discordance.^76^ Those with limited English language proficiency have been shown to have lower healthcare access^72^ as well as worse blood pressure control^77^ and health outcomes^78, 79^ compared to those with proficient English language skills. Thus, to reduces disparities in sodium reduction advice and reach the most at-risk populations, the health system should prioritize recruitment, retention, and training of a diverse healthcare workforce,^81^ increasing the availability of medical interpreters,^80^ and the provision of culturally and linguistically appropriate educational materials.

After reducing processed food consumption, the most common physician advice for sodium reduction is to read nutrition labels.^70^ Nutrition label requirements apply to all domestic and imported foods in the US.^82^ Despite the ubiquitousness of these labels, across racial and ethnic groups 28% to 43% of adults rarely or never use these labels. Although label reading has been associated with better dietary intake,^83, 84^ nutrition fact panels, such as those on the back of products in the US, are more difficult to interpret than front of pack (FOP) labels, especially compared to those that aid interpretation through visuals (e.g., traffic lights).^85–87^ Such visuals may help to overcome language and health literacy barriers that may limit the use of the current labels. As such the World Health Organization recommends countries adopt FOP labelling schemes^14,81^ and the FDA is currently considering FOP labels.^81, 88^

To our knowledge, this is the first report to examine how the underlying assumption that rice is prepared using salt affects sodium estimates. The practice of salting or flavoring plain rice may vary across cultures. Notably, two prior assessments of sodium sources using NHANES data also mention this assumption as a potential limitation^31, 32^ given rice has not been shown to be a major source of sodium in China or Japan.^89, 90^

Analyses of NHANES data have consistently shown Asian Americans have the highest sodium intake of all racial and ethnic groups based on 24-hr dietary recalls.^4, 19, 30, 32, 91–93^ The ∼325 mg/day difference in sodium estimates observed when varying the assumption that rice is salted is enough to shift Asian Americans from being amongst those with the highest intake to being amongst those with the lowest intake. This finding reinforces calls to challenge dietary research measures, concepts, definitions, and dietary guidance to promote inclusivity^94, 95^ as well as the need for more nuanced Asian American health and nutrition research.^96–98^

Given our results, Asian American sodium appears to be similar relative to other racial and ethnic groups. This aligns with data from both a representative sample of New York City adults and the Multiethnic Study of Atherosclerosis (MESA) which measured sodium intake via 24-hour and spot urine, respectively, and found no difference between Asian American sodium intake and that of other racial and ethnic groups.^99, 100^ More research is needed to understand differences between Asian American subgroups (e.g., although data from the Mediators of Atherosclerosis in South Asians Living in America study and MESA suggest no differences between Chinese and Indian Americans)^101^ as well as differences by acculturation (e.g., as data from the Multiethnic Cohort, suggest differences by nativity).^102^

Finally, the National Strategy on Hunger, Nutrition, and Health recommends that data collection be improved to better identify trends in sodium intake.^81^ Thus, the most important take-away of this analysis is that the FNDDS database and ASA-24 should be updated to allow users to choose salted or unsalted rice as in other dietary data systems (Caitlin Caspi, ScD, email communication, June 13, 2024).^104–106^ Because 24-hr recall underestimates sodium compared to 24-hr urine,^107^ 24-hr urine is recommended to describe sodium intake and monitor sodium trends.^108, 109^ Thus, it also may be necessary to regularly collect 24-hr urine in a subsample of NHANES participants.

### Strengths and Limitations

A strength of this analysis is the provision of racial and ethnic specific estimates for both sodium sources and sodium reduction behaviors, and in particular, the disaggregation of “Hispanic” into Mexican Americans and Other Hispanic adults. This disaggregation revealed both similarities and differences between the groups in sodium sources and behaviors.

However, no disaggregation of the Asian American or the Other Race categories was possible with the NHANES public use dataset. As NHANES sampling of Asian Americans, Hispanics, and Other Race groups is not intended to be representative of the subpopulations within these groups, our results may fail to reflect the cultural, linguistic, and economic diversity present. This inability to disaggregate could easily obscure important differences. For example, some of the top sodium sources for Asian Americans (e.g., soy-based condiments, stir-fry and soy-based sauce mixtures) are more representative of an East and Southeast than South Asian dietary pattern.^110^ As both the Asian American and multiracial populations are growing, more attention will need to be paid to the health needs and health messaging of these groups.^20, 111^

Additional limitations of this analysis relate to the complexity of measuring long-term dietary intake with a short-term instrument such as a dietary recall.^112^ Although sodium intake is underestimated using dietary recalls,^107^ population level estimates are still reasonable approximations of intake. Similar analyses may produce varying results which may not be attributable to temporal changes in eating patterns or the population under study but, as already noted, may be due to underlying assumptions of food and nutrient databases or the grouping of foods into categories. Finally, self-reported engagement in sodium reduction behaviors may be inflated due to social desirability bias.

## Conclusion

Racial and ethnic disparities in blood pressure awareness and control^15^ might be reduced by adopting various sodium reduction strategies, which can be informed by observed differences in the top sources of sodium and sodium reduction behaviors. Cultural tailoring might be used for individual-level educational interventions in primary care or the community and in public health messaging. Promoting discretionary salt substitution may be a viable strategy among some subgroups of the population in addition to being used in product reformulation. Given the reportedly low use of nutrition facts labels, efforts to aid interpretation would be warranted through front-of-pack labelling schemes. Sodium reduction initiatives at multiple levels,^87^ especially changes to the food supply through mandatory and/or voluntary sodium reduction targets, will be needed to reduce sodium to recommended levels. Finally, current dietary data collection methods and concepts may need to be challenged in order to promote inclusivity of diverse eating patterns.^94^

## Data Availability

The analytical code used to generate results is publicly available and can be accessed at Open Science Framework (DOI 10.17605/OSF.IO/ER4V9). JC had full access to all the data and takes responsibility for its integrity and the data analysis.

https://osf.io/er4v9/

## Acknowledgements

None

## Supplemental Material

Tables S1-2

Figure S1

---

**Supplemental Table 1.**
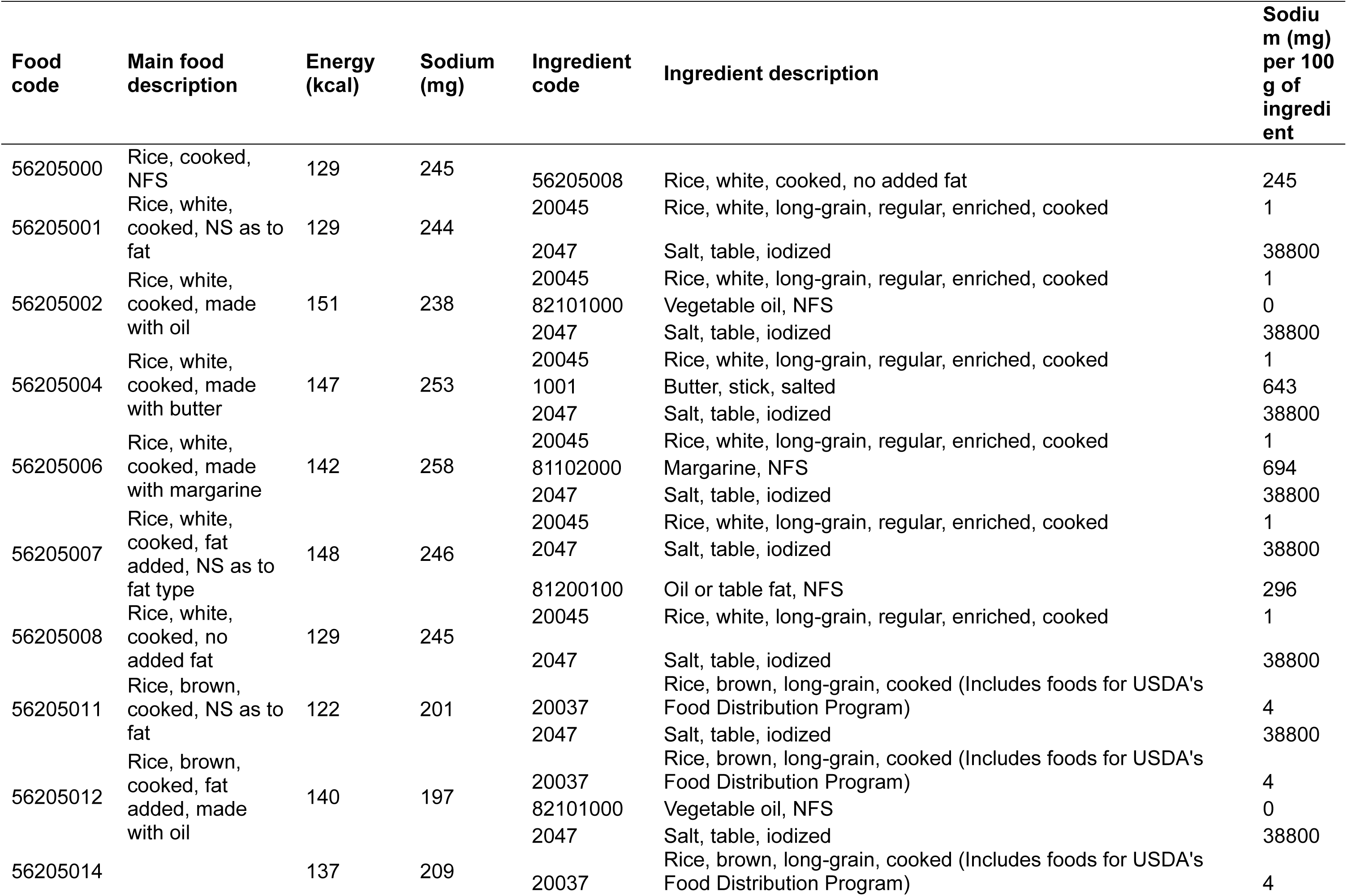
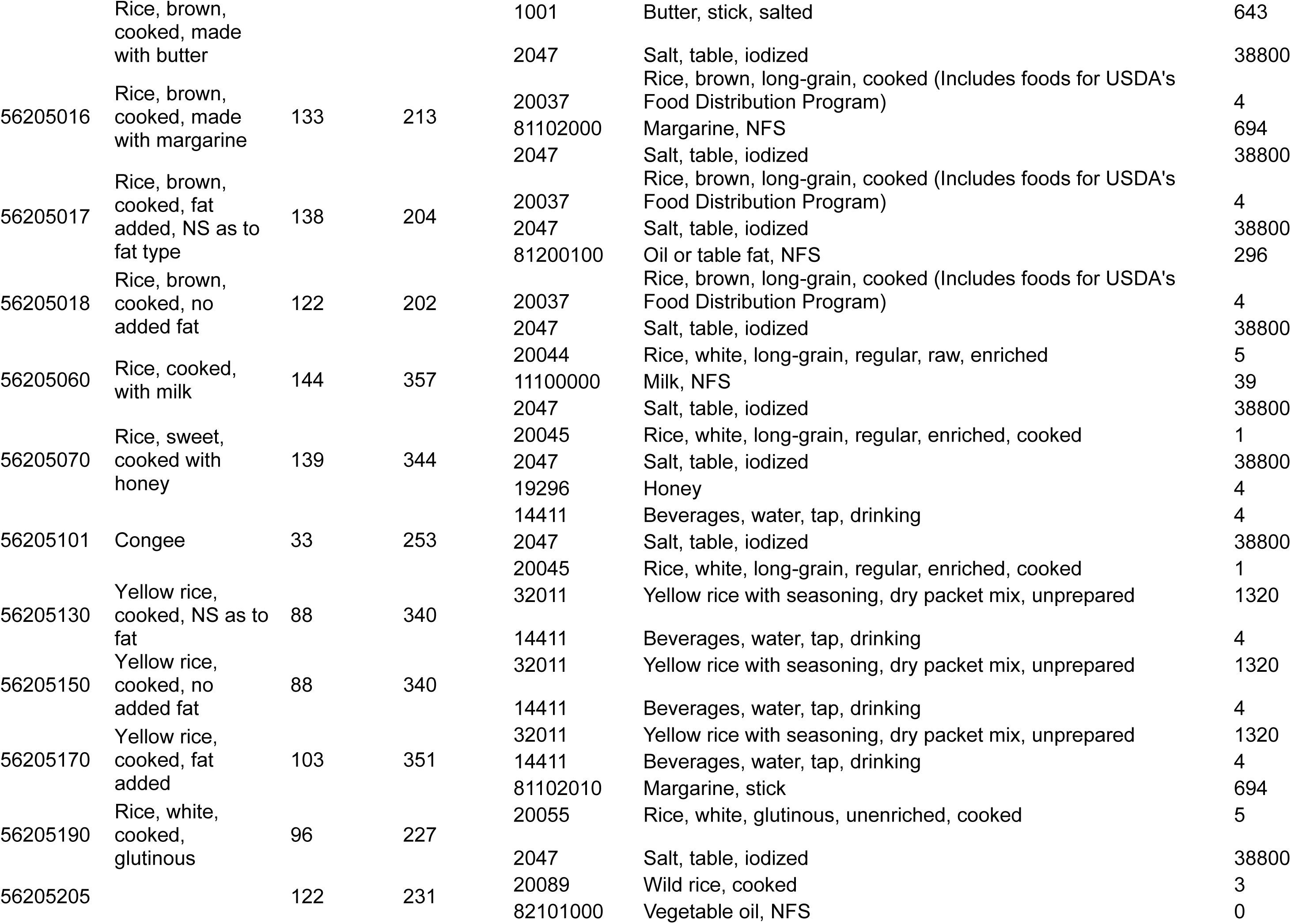
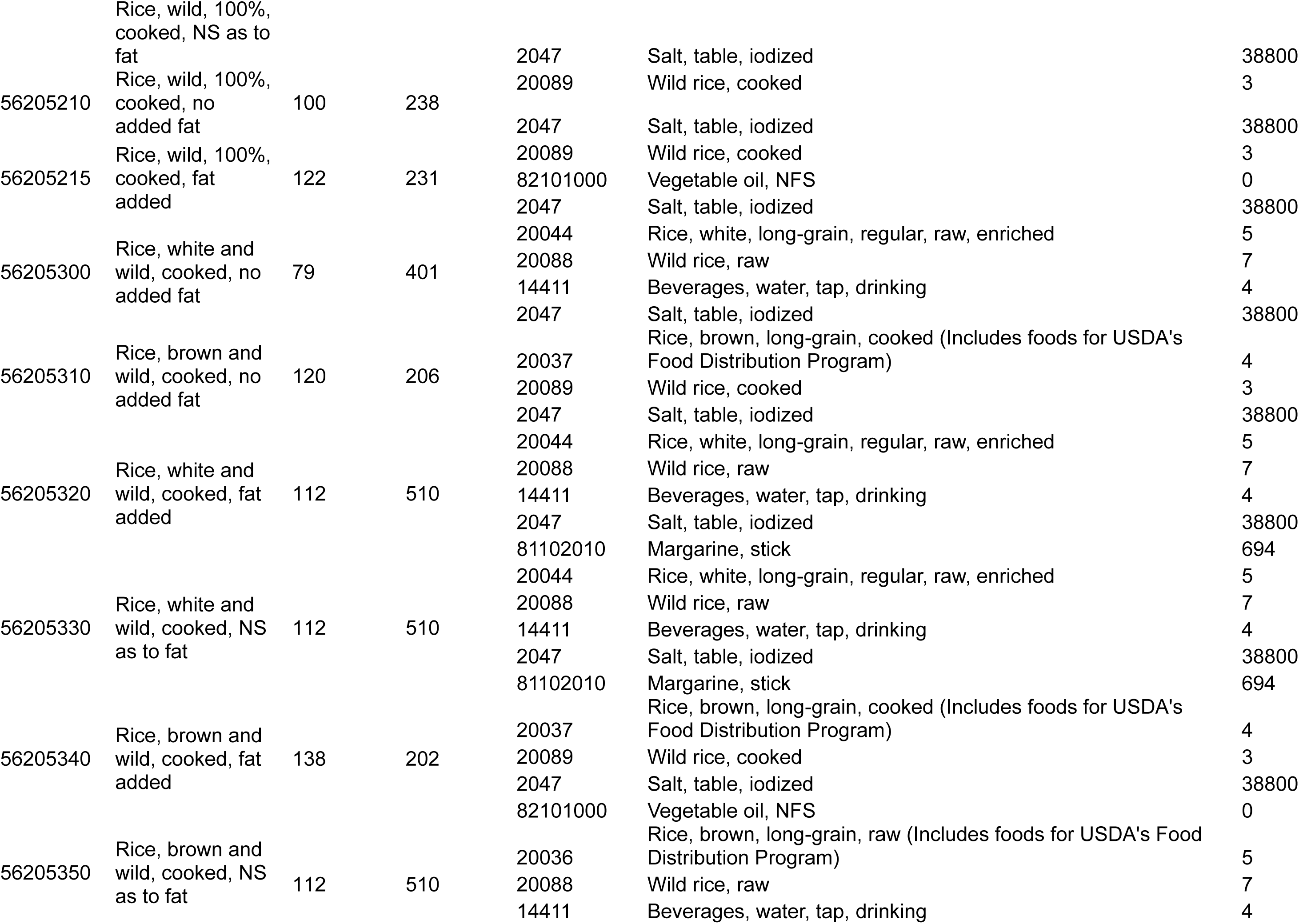
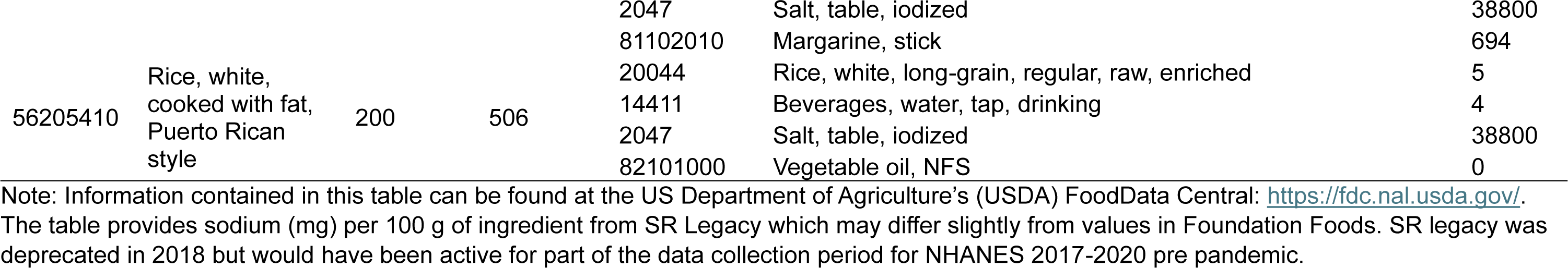
Sodium Content and Ingredients of What We Eat in America (WWEIA) Food Codes in the “Rice” Category.

**Supplemental Table 2:** US Adult Sodium Intake (mg/day) and Percent of Population with Excessive Sodium Intake under Varying mptions about Salt in Rice by Race and Ethnicity, NHANES 2017-2020 pre-pandemic.

| **Salt Assumption** | **Race** | **Unweighted N** | **Mean (SE)** | **25<sup>th</sup> Percentile (SE)** | **Median (SE)** | **75<sup>th</sup> Percentile (SE)** | **Excessive Percent (SE)\*** |
| --- | --- | --- | --- | --- | --- | --- | --- |
| **Salt in Rice<sup>†,‡</sup>** | **Mexican American** | 952 | 3675 (63.9) | 2914 (60.6) | 3576 (63.5) | 4329 (74.8) | 91.9 (1.2) |
|  | **Other Hispanic** | 820 | 3232 (92.5) | 2526 (69.9) | 3129 (90.8) | 3825 (114.3) | 83.1 (2.2) |
|  | **Asian American** | 877 | 3500 (60.7) | 2758 (47.2) | 3400 (59.6) | 4126 (76.0) | 89.0 (1.2) |
|  | **Black** | 2153 | 3267 (45.5) | 2562 (44.8) | 3167 (46.8) | 3869 (54.6) | 84.1 (1.4) |
|  | **Other Race** | 397 | 3606 (97.2) | 2852 (89.6) | 3512 (97.9) | 4251 (109.6) | 90.8 (1.9) |
|  | **White** | 2893 | 3467 (41.5) | 2733 (46.4) | 3370 (42.9) | 4089 (46.3) | 88.4 (1.3) |
| **No Salt in Rice<sup>†,§</sup>** | **Mexican American** | 952 | 3632 (64.6) | 2873 (60.7) | 3532 (64.3) | 4285 (75.5) | 91.2 (1.3) |
|  | **Other Hispanic** | 820 | 3114 (97.7) | 2421 (75.8) | 3011 (96.6) | 3695 (118.9) | 79.5 (2.7) |
|  | **Asian American** | 877 | 3177 (58.2) | 2476 (45.1) | 3079 (56.3) | 3764 (72.8) | 81.4 (1.6) |
|  | **Black** | 2153 | 3222 (45.3) | 2519 (42.1) | 3122 (46.0) | 3821 (55.4) | 82.8 (1.4) |
|  | **Other Race** | 397 | 3545 (91.8) | 2794 (84.3) | 3451 (92.2) | 4185 (103.5) | 89.7 (2.0) |
|  | **White** | 2893 | 3441 (42.4) | 2706 (44.5) | 3342 (43.0) | 4062 (48.8) | 87.8 (1.2) |

\*Excessive Percent refers to the percent of the population estimated to have sodium intake above the 2020-2025 Dietary Guidelines for Americans recommended limit of 2300 mg/day.
† "Rice" includes food codes: 56205000, 56205001, 56205002, 56205004, 56205006, 56205007, 56205008, 56205011, 56205012, 56205014, 56205016, 56205017, 56205018, 56205060, 56205070, 56205101, 56205130, 56205150, 56205170, 56205190, 56205205, 56205210, 56205215, 56205300, 56205310, 56205320, 56205330, 56205340, 56205350, and 56205410.
‡Salt in Rice: The sodium content was not altered for any of the food codes in the "rice" category.
§No salt in Rice: Sodium due to salt was removed from all foods codes in the "rice" category except for yellow rice (foods codes: 56205130, 56205150, and 56205170).

**Supplemental Figure 1:**
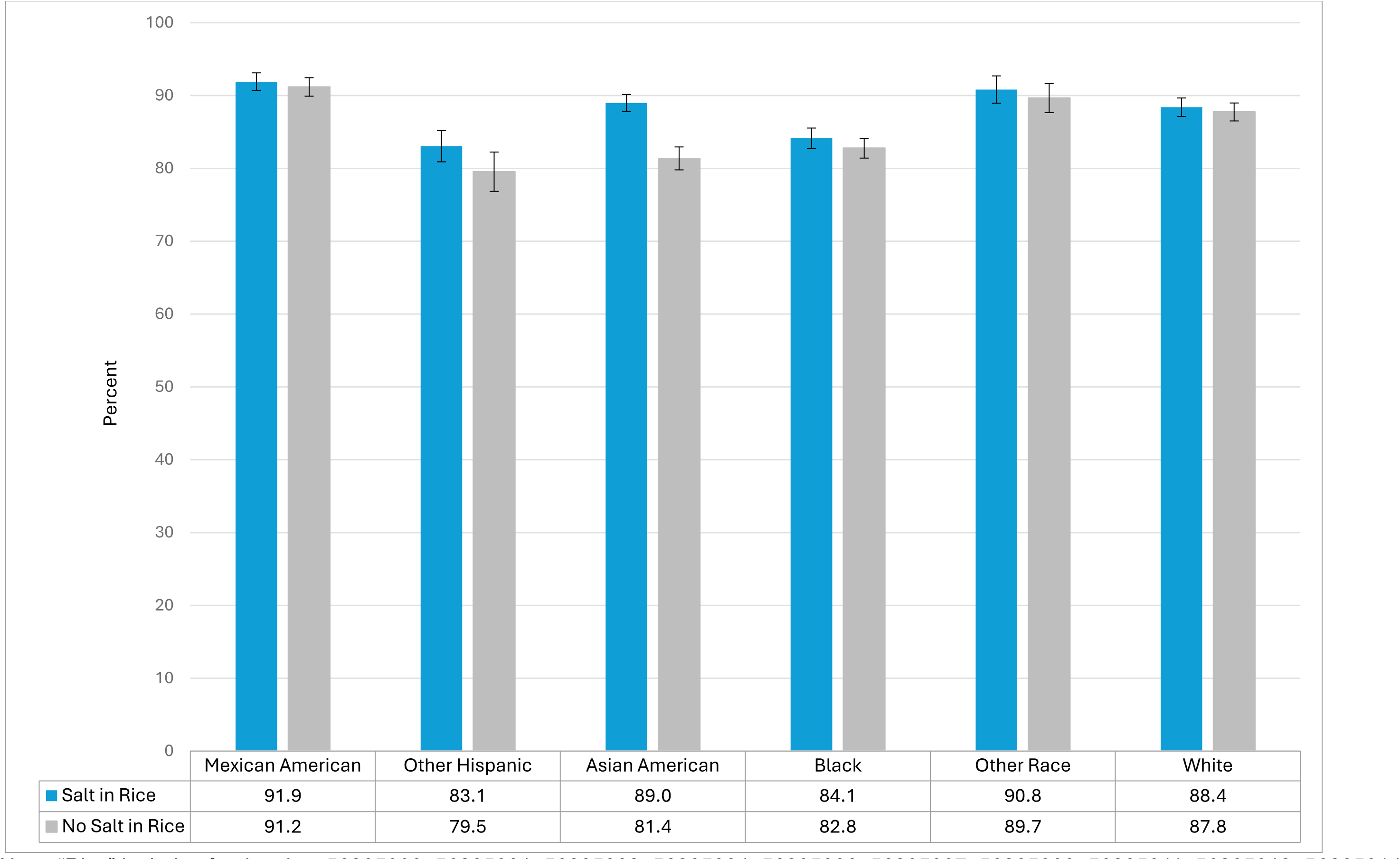
Percent of the US Adult Population above Recommended 2300 mg/day of Sodium Intake and Standard Error by & Ethnicity under Varying Assumptions about Salt in Rice, NHANES 2017-2020 pre-pandemic. Note: “Rice” includes food codes: 56205000, 56205001, 56205002, 56205004, 56205006, 56205007, 56205008, 56205011, 56205012, 56205014, 56205016, 56205017, 56205018, 56205060, 56205070, 56205101, 56205130, 56205150, 56205170, 56205190, 56205205, 56205210, 56205215, 56205300, 56205310, 56205320, 56205330, 56205340, 56205350, and 56205410.

